# A systematic review of genome-wide association studies on bladder cancer

**DOI:** 10.1101/2023.09.17.23295670

**Authors:** Aliyu Adamu Ahmad, Umar Muhammad, Buhari Ibrahim, Suleiman Hamidu Kwairanga, Usman Adamu Garkuwa, Murtala Muhammad Jabril, Umar Ahmad

## Abstract

**Background:** Bladder cancer (BC) is the most common cancer of the urinary tract worldwide with over 550,000 new cases each year, bladder cancer has drawn relatively limited research attention and healthcare interventions despite the escalating incidence and mortality rates, particularly in Africa. Historically, the clinical handling of bladder cancer remained largely unchanged for many years. However, novel research initiatives have heralded a fresh epoch in its diagnosis and treatment, fueled by detailed probing of molecular changes.

**Aim:** This study aimed to identify genetic susceptibility loci associated with bladder cancer by systematically reviewing previous Genome-Wide Association Studies (GWAS).

**Methods:** In line with this objective, comprehensive literature searches were conducted across PubMed, Google Scholar, and relevant genetic databases, focusing on bladder cancer GWAS studies from 2000 through to November 2022. This systematic review adhered to the robust PRISMA standards. To evaluate the credibility of the studies under scrutiny, the Newcastle-Ottawa Scale was employed, further assessing any potential bias risk.

**Results:** The investigation identified chromosome 18q12.3 as the most vulnerable to bladder cancer, revealing four polymorphisms at this locus: rs7238033, rs10775480, rs11082469, and rs17674580. Furthermore, chromosome 5p15.3 emerged as the second most susceptible, with three noted polymorphisms: rs2736098 and two instances of rs401681.

**Conclusion:** Despite these findings, our understanding of genetic predisposition to bladder cancer remains rudimentary, with the majority of substantial data deriving from GWAS. No additional genetic association evidence emerged from this systematic review. Given the relatively minor influence of our current knowledge of genetic susceptibility to bladder cancer on public health, a call for larger cohort studies is necessary. These expanded studies can potentially unveil a broader range of significant polymorphisms across the genome, thereby enhancing our understanding and approach to bladder cancer.

## 1.0 Introduction

Bladder cancer, a multifaceted disease originating from the confluence of genetic and environmental risk factors, ranks as the sixth most prevalent cancer type worldwide. Annually, it accounts for approximately 150,300 fatalities (Selvaraj et al., 2021; Wang *et al*., 2016). Both inherited genetic factors and environmental aspects like smoking and occupational exposure stand as major risk factors for bladder cancer (Wang *et al* ., 2016).Moreover, other elements such as lifestyle, medical history, fluid intake, and diet also contribute to its neogenesis(Wang *et al* ., 2016).

In the United States alone, an assessment in 2015 reported that bladder cancer affected 56,329 males and 17,680 females, resulting in 16,000 deaths (Nahar et al., 2011). Prevalence of bladder cancer risk is notable among Asian populations (Bau et al., 2011)(Wang et al., 2016). However, the exact factors and genes leading to this disease are yet to be fully identified. Despite most patients with bladder cancer undergoing conservative surgery, they face an exceedingly high risk of frequent recurrences. This aspect escalates the healthcare cost, making bladder cancer one of the most expensive cancers to manage in many Western countries(Nahar et al., 2011; Ye et al., 2011).

Interestingly, in Africa, bladder cancer incidence and mortality rates have been on an alarming upward trajectory. Despite the scarcity of comprehensive data compared to Asian and Western demographics, some regions in Africa report substantially high bladder cancer incidence rates, pointing to an emerging public health issue (WHO, 2022). Alarmingly, research and interventions for bladder cancer in Africa have been insufficient, further highlighted by the paucity of genetic studies and adequate healthcare services for early diagnosis and treatment. Genome wide associated studies (GWAS) is a study designed to collect and compare different type of genes in order to identify genes or genetic variant that may contribute to a particular type of disease among populations (Chang et al., 2018). Genetic polymorphism is a phenomenon in which an individual’s genes have a variety of DNA sequences. The polymorphisms of the DNA sequences, which may vary by two or more, have a role in determining the diversity of people, groups, and populations. Single nucleotide polymorphism (SNP) analyses the human genome for variations that are more prevalent in people with a certain type of disease (Verma, 2016).

In this study, a systematic reviews approach were employed to thoroughly search and identify all important published studies on bladder cancer and GWAS. The is aims to identify genetic susceptibility loci associated with bladder cancer by systematically reviewing previous GWAS studies. This approach will help further our understanding of genetic predisposition to bladder cancer, ultimately leading to enhanced preventative measures and treatment strategies.

## 2.0 Materials and methods

### 2.1 Protocol and registration

The Preferred Reporting standards for Systematic Reviews and Meta-Analyses (PRISMA) served as our guide for conducting this systematic review (Shamseer *et al*., 2015). The review protocol was registered with PROSPERO and the study ID; CRD42022351101 is obtained (https://www.crd.york.ac.uk/prospero/display_record.php?ID=CRD42022351101).

### 2.2 Selection criteria

The inclusion criteria for this systematic review were as follows: studies had to be published in peer-reviewed journals and be written in English. The design of the selected studies should be specifically GWAS with a focus on identifying intriguing genetic variants, particularly SNPs, related to bladder cancer. No restrictions were made based on language, demographics of the patients, or their ages. However, review is also excluded to prevent duplication of data.

### 2.3 Search strategy and data source

Articles publish for the time period between January 1, 2000 and November 17, 2022, were retrieved from PubMed, the Cochrane Database of Systematic Reviews, ScienceDirect and the Lippincott database respectively. The searched databases, date, time of last search, search strategy employed, and the total number of results obtained in each database are described in in Table 1. Search strategies and search terms are in supplementary 1.

**Table 1:** Characteristics of the thirteen (13) included studies.

| S/<br>No. | Database | Date and time<br>of last search | Type of<br>search strategy<br>employed | Num<br>ber of<br>articles<br>found |
| --- | --- | --- | --- | --- |
| 1 | PubMed (pubmed.ncbi.nlm.nih.gov/) | 13 November 2022<br>11:34 | MeSH | 157 |
| 2 | Google Scholar (scholar.google.com/) | 18 November 2022<br>09:20 | Boolean logic | 18,000 |
| 4 | ScienceDirect<br>(www.sciencedirect.com/) | 14 November 2022<br>18:57 | Advance search | 16,972 |
| 5 | Cochrane Library<br>(www.cochranelibrary.com/) | 14 November 2022<br>19:02 | Key terms | 829 |
| 6 | Lippincott<br>(https://lww.com/) | 15 November 2022<br>7:56 | Boolean logic | 866 |

Our chosen search terms were tailored to suit the unique attributes of each database. For additional studies, we scrutinized the bibliographies of the studies included in our initial search, as well as the references in ISI Web of Knowledge. Our search strategy centered around keywords pertinent to Genome-Wide Association Studies (GWAS), bladder cancer, and susceptibility genes, which we merged with the MESH terms. To facilitate the review process, we employed Covidence, an online tool designed to streamline the screening and data extraction stages of systematic reviews (Kellermeyer *et al*., 2018). Two independent co-reviewers, Aliyu Adamu Ahmad and Umar Muhammad separately screened the downloaded articles. The screening was based on the article title, the defined keywords, and the abstract, in line with the pre-established inclusion criteria.

### 2.4 Study selection

Upon concluding the title and abstract screening, we gathered all potentially relevant full-text reports and publications. Two independent reviewers performed a thorough assessment of these full texts. Any studies that were excluded at this stage were documented along with the reasons for their exclusion. In the event of disagreements, resolution was reached through discussion among the review team members. We have provided a detailed account of the number of records retrieved from the literature searches, the quantity of included and excluded studies, as well as the specific reasons for exclusions. This information is represented via a PRISMA flow diagram, as illustrated in Figure 1.

**Figure 1:**
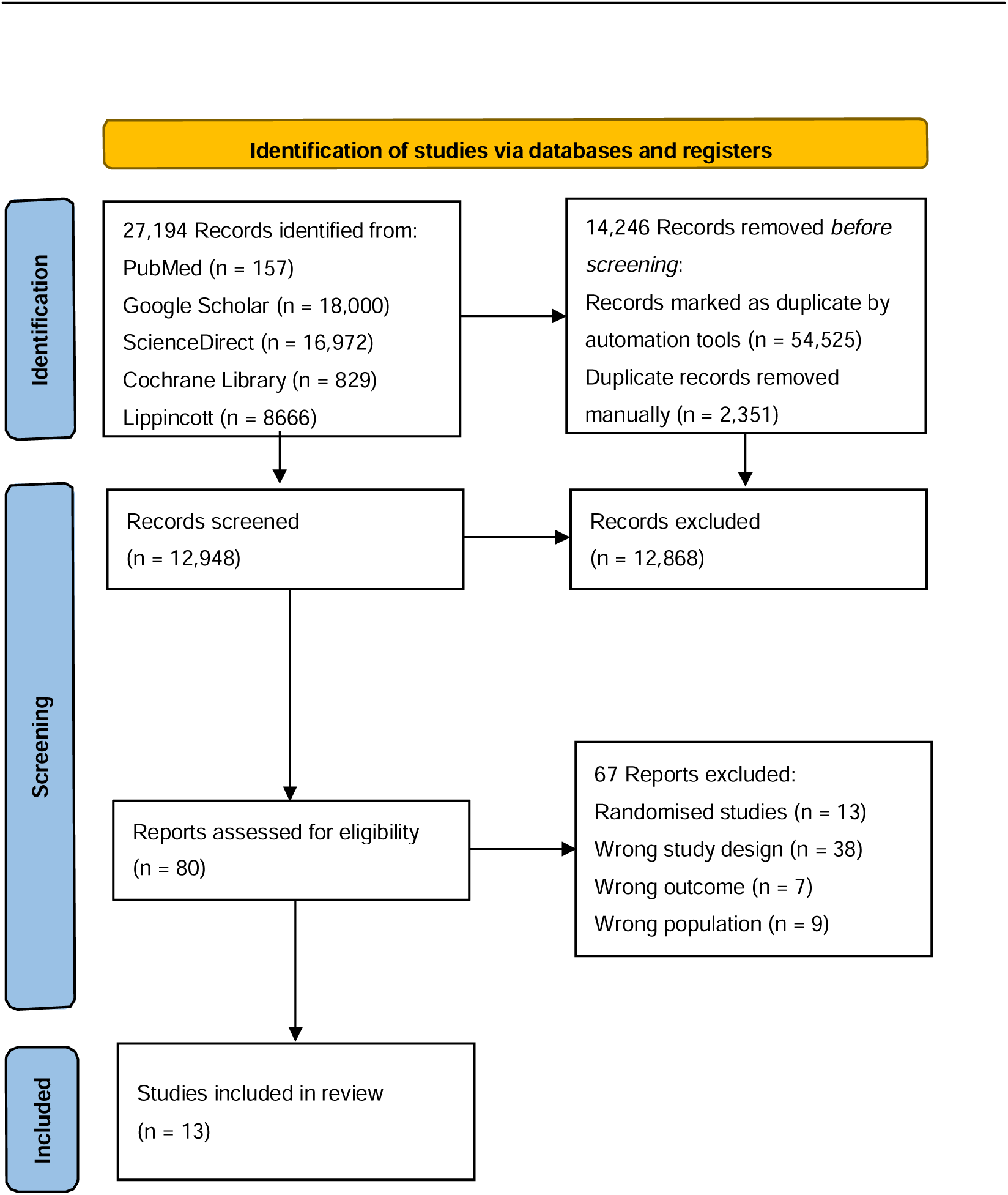
PRISMA Graphic: Indicating the process of identifying and choosing the studies included.

### 2.5 Data extraction

A customized data extraction sheet was used to capture information about; the first authors and year of publication, country of study, study design, case and controls, methods, genetic variants (SNPs), locus, genes, alleles, *p* values and associations. This step was carried out by the first reviewer (Aliyu Adamu Ahmad) and the extracted data were then cross-checked by the second reviewer (Umar Muhammad).

### 2.6 Quality assessment

Two reviewers (AAA and UM) independently assessed the quality of each included article using the Newcastle-Ottawa Scale (NOS) (Lo *et al*., 2014). The NOS is made out of eight components that are divided into three domains: (1) choosing study groups, (2) determining exposure and results, and (3) group comparability to gauge the caliber of observational studies. Studies received a maximum rating of nine stars, and ratings were based on a star system. Studies with less than three stars were labelled as low quality, those with four to six stars as moderate quality, and those with seven to nine stars as high quality. In order to minimize errors and reduce bias, steps in the review process were independently duplicated, especially when choosing studies and extracting data. Independent reviewers looked at both eligible and ineligible articles after the initial screening of the articles.

## 3.0 Results

### 3.1 Literature search

Figure 1 depicts the flow diagram outlining the procedures involved in article selection in accordance with PRISMA standard for systematic review (Shamseer *et al*., 2015). The search strategy generated 16,972 records after removing the duplicates. Upon collating the search results from the six electronic databases (PubMed, Google Scholar, ScienceDirect, Cochrane Library, and Lippincott), a total of 12,868 studies were ruled out. The reasons for disqualification were their irrelevance to Genome-Wide Association Studies (GWAS), lack of focus on bladder cancer in human subjects, or absence of an abstract.

Following the screening of titles and abstracts, we undertook a detailed examination of the full texts of the remaining 80 articles, evaluated against the predetermined inclusion criteria. At this stage, studies were excluded for several reasons: randomized study design (n = 13 instances), inappropriate study design (n = 38 instances), unsuitable setting (n = 9 instances), incorrect outcome focus (n = 6 instances), and inappropriate population (n = 1 instance). Upon thorough evaluation and application of the selection criteria, thirteen studies were ultimately deemed suitable for inclusion in this review. These selected studies originated from various authors and were published between the years 2009 and 2022. The comprehensive list includes studies conducted by Figueroa *et al*., (2015), Gu *et al*., (2011), Matsuda *et al*., (2015), Garcia-closas *et al*., (2011), Wang *et al*., (2009), Rafnar *et al*., (2011), Wu *et al*., (2012), Figueroa *et al*., (2014), Matsuda *et al*., (2015), Kiemeney *et al*., (2010), (Xu *et al*., 2020), Mamdouh *et al*., (2022), and Stern *et al*., (2009).

### 3.2 Characteristics of the data extracted from the studies

Table 1 summarizes the main characteristics of the selected studies that identified the genetic susceptibility loci associated with bladder cancer by systematically reviewing previous genome-wide association studies (GWAS). All included studies had a case–control study design (Figure 2A). Most of these studies (62%) had ≥ 1000 participants in both the case and control groups. Six studies (46%) were conducted in America, three studies (23%) in Asia, Europe had two studies (15%), one study in Africa (8%) and, interestingly, one study (8%) was jointly conducted in America and Asia (China and USA) (Figure 2B & C). Furthermore, out of the 22 genetic susceptibility loci associated with bladder cancer identified in the included studies, the most frequently appearing SNP was 20p22.2 (seven times), followed by 18q12.3 (four times), and 3q28 and 5q15.3 appeared three times each (Figure 3A). For allele, eight different combinations were observed, with C/T appearing most frequently (nine times) and A/T appearing less frequently (one time) (Figure 3B). In addition, rs6104690, rs71052, and rs9642880 were the most appearing SNPs (appeared three times each) and rs401681 and rs1014971(appeared twice each) (Figure C). For genes, PSCA appeared more often than any other gene.

**Table 1:** Records of the databases consulted, date and time of the last search, the search strategy employed and the number of records found.

| S/No | Author (year) | Country of study | Case and controls (n) | Genotype (SNPs) | Locus | Gene | Allele | P value | Association |
| --- | --- | --- | --- | --- | --- | --- | --- | --- | --- |
| 1 | Jonine D. Figueroa (2016) | USA | Case (5551)<br>Control (10242) | rs6104690<br>rs6108803<br>rs62185668 | 20p12.2<br>20p12.2<br>20p12.2 | | A/G<br>G/A<br>A/C | 5.27E-03<br>3.33E-02<br>2.93E-03 | rs6104690 and rs6108803 exhibited significant links with bladder cancer risk ($P \leq 0.03$) regardless of the models, when accounting for the recently discovered 20p12.2 SNP rs6108803, the link for rs62185668 was no longer significant in ($P = 0.25$). |
| 2 | Jian Gu (2011) | USA | Case (969)<br>Control (946) | rs602846<br>rs621559<br>rs398652<br>rs654128 | 20q11.22<br>1p34.2<br>14q21<br>6q22.1 | *PELI2* | N/A<br>G/A<br>G/A<br>C/A | 0.024833<br>0.000364<br>0.028771<br>0.017219 | the SNP rs398652 on 14q21 and numerous other intriguing genetic variations were linked to leukocyte telomere length, and rs398652 is discovered that it also links to a lower risk of bladder cancer. Increased telomere length was linked to the variant alleles of all four of these SNPs ($P < 105$) |
| 3 | Matsuda K. (2015) | Japan | Case (1131)<br>Control (12558) | rs11543198 | 15q24 | *CYP1A2* | G/A | 1.22E-102<br>4 | Chromosome 15q24's single SNP, rs11543198, showed a significant correlation with a P-value of 1.22 1024. |
| 4 | Montserrat Garcia-Closas (2011) | USA | Case (5883)<br>Control (8277) | rs7238033<br>rs10775480<br>rs11082469 | 18q12.3<br>18q12.3<br>18q12.3 | *SLC14A1* | C/T<br>C/T<br>A/G | 8.72E-09<br>8.95E-09<br>1.84E-05 | The three SNPs in the locus 18q12.3 revealed risk associations with BCa and Similar signals were also seen for rs7238033 and rs10775480/rs10853535. |
| 5 | Meilin Wang (2009) | China | Case (415)<br>Control (464) | rs9642880<br>rs710521 | 8q24<br>3q28 | *MYC* | G/T<br>A/G |  | The rs710521 A/G polymorphism, which was linked to a higher risk of bladder cancer, is absent, but rs9642880 G/T is present. |
| 6 | Thorunn Rafnar (2011) | Iceland | Case (4147)<br>Control (34988) | rs2736098<br>rs401681 | 5p15.33<br>5p15.33 | *CLPTM1L*<br>*PSCA* | G/A<br>C/T | 0.125<br>0.254 | examination of the rs2736098 (A) region of the TERT- CLPTM1L gene, which indicated a greater relationship with specific cancer types. However, neither variation could entirely explain the rs40168 association with bladder cancer. |
| 7 | Xifeng Wu (2009) | USA | Case (969)<br>Control (957) | rs2294008 | 8q24.3 | *PSCA* | C/T | 0.010 | In US and European populations, a missense mutation (rs2294008) in the PSCA gene consistently correlated with bladder cancer. |
| 8 | Nathaniel Rothman (2010) | USA | Case (3532)<br>Control (5120) | rs1014971<br>rs8102137<br>rs11892031<br>rs1495741 | 22q13.1<br>19q12<br>2q37.1<br>8p22 | *CBX6*<br>*CCNE1*<br>*UGT1A*<br>*NAT2* | T/C<br>T/C<br>A/C<br>A/G | 0.001<br>0.485<br>0.002<br>0.188 | This study discovered three novel areas, rs1014971, rs8102137, and rs11892031, on chromosomes 22q13.1, 19q12, and 2q37.1 which are linked to bladder cancer. |
| 9 | Jonine D. Figueroa (2013) | USA | Case (2422)<br>Control (5751) | rs10936599<br>rs907611<br>rs6104690<br>rs4510656 | 3q26.2<br>11p15.5<br>20p12.2<br>6p22.3 | *MYNN*<br>*LSP1* | T/C<br>G/A<br>C/A<br>G/A | 4.53E-9<br>4.11E-8<br>7.13E-7<br>6.98E-7 | Following normal quality control, it was determined that the following SNPs: rs10936599, rs907611, rs6104690, and rs4510656 have bladder cancer risk associations. |
| 10 | Chenyang Xu (2020) | China | Case (581)<br>Control (1561) | rs401681<br>rs9642880<br>rs4907479<br>rs17674580<br>rs4813953<br>rs6104690<br>rs1014971 | 5p15.3<br>8q24.21<br>13q34<br>18q12.3<br>20p12.2<br>20p12.2<br>22q13.1 | *CLPTMIL*<br>*CASC11*<br>*MCF2L*<br>*SLC14A1*<br>*LOC33959*<br>*3*<br>*C20orf187*<br>*APOBEC3*<br>*A* | C/T<br>C/T<br>A/G<br>C/T<br>C/T<br>A/G<br>C/T |  | Among the eight SNPs examined in this investigation, the Chinese population showed an association with BCa risk at rs798766 at 4p16.3, rs9642880 at 18q12.3, and rs4813953 at 20p12.2. |
| 11 | Samah Mamdouh (2021) | Egypt | Case (150)<br>Control (50) | rs9642880<br>rs710521 | 8q24.21<br>3q28 | *MYC*<br>*TP63* | G/T<br>A/G |  | rs9642880 G/T, and rs710521 A/G are both identified a and substantially related with the risk of bladder cancer. |
| 12 | Mariana C. Stern (2009) | USA/China | Case (1042)<br>Control (1123) | rs710521 | 3q28 | *TP63* | A/T | 0.080 | Our findings imply that the rs710521 A allele might raise the risk of developing urinary low risk bladder tumours. |
| 13 | Lambertus A Kiemeneij (2010) | Netherlands | Case (4,739)<br>Controls 45,549 | rs798766 | 4p16.3 | *TACC3* |  | 9.9E-12 | UBC was found to be associated with the T allele of rs798766 on 4p16.3 (P = 9.9 1012). |

**Figure 2:**
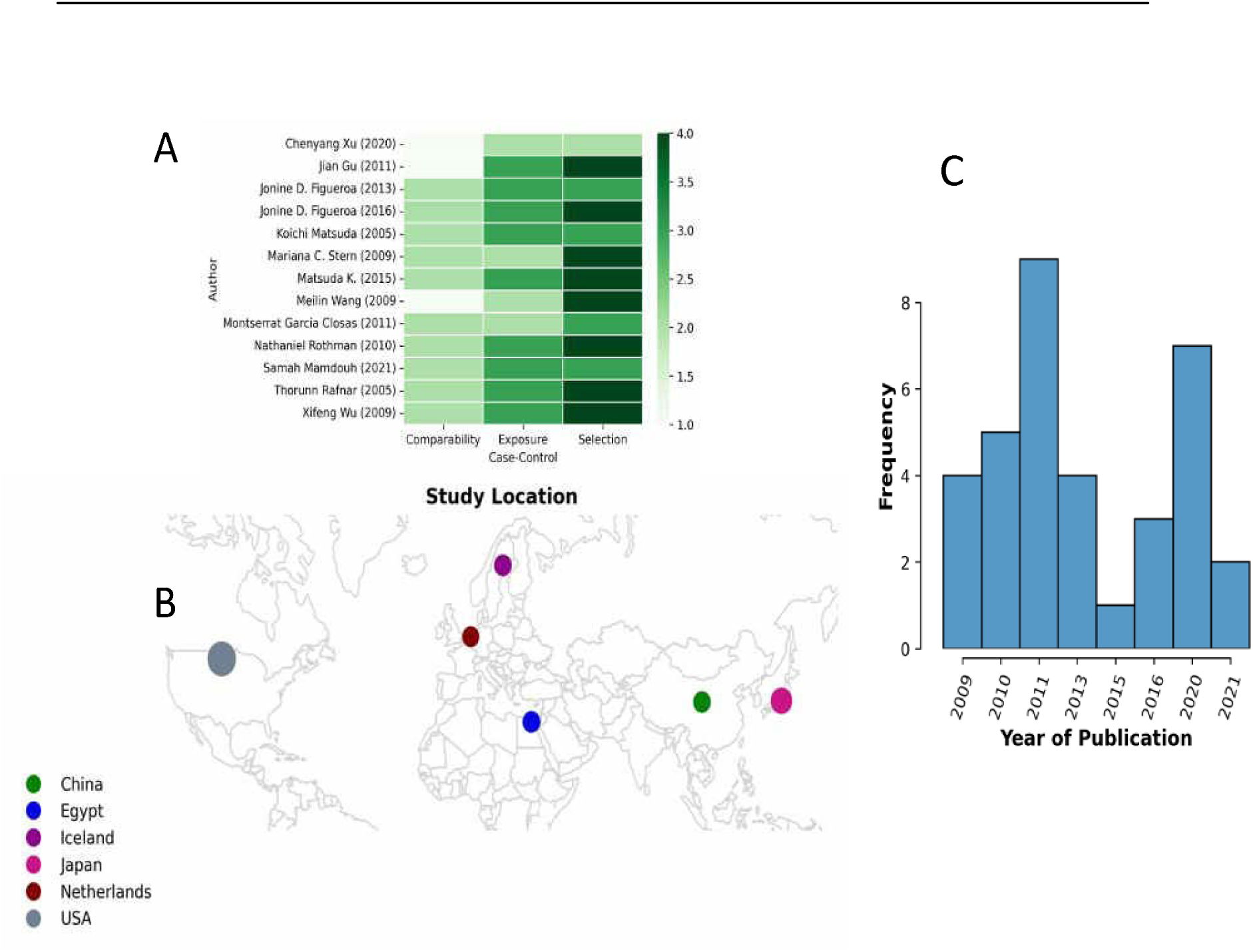
Publication features. A) All the included articles are case-control designed studies., B) Map of the study locations, showing countries where the studies were carried out. Coloured-circle are according to the number of papers per each country., C) Frequency of studies per year of publication.

**Figure 3:**
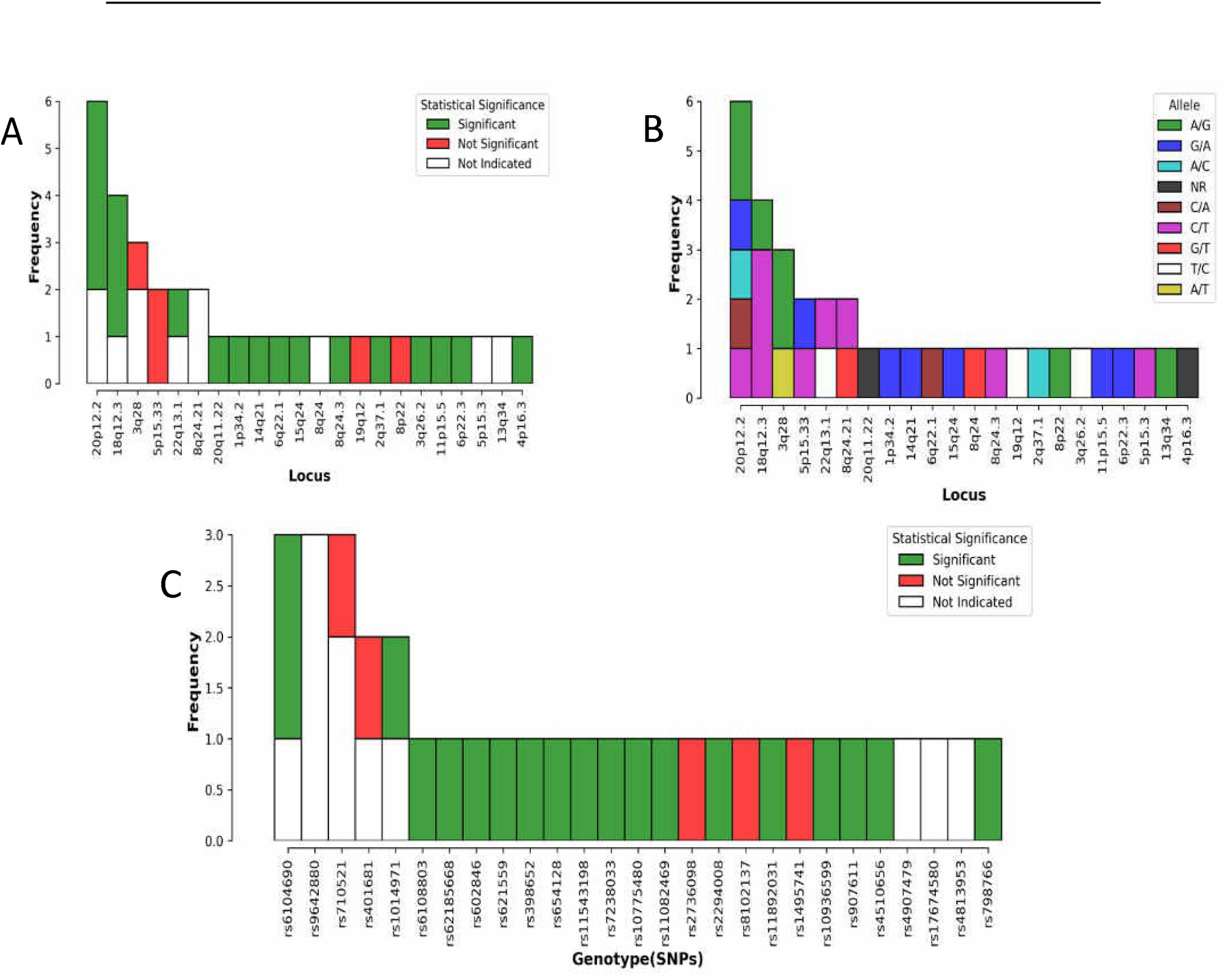
Features of the extracted GWAS data. A) Frequency of 22 genetic susceptibility loci associated with bladder cancer identified from the studies., B) Allele of frequency., C) Frequency of genotype (SNPs).

**Figure 4:**
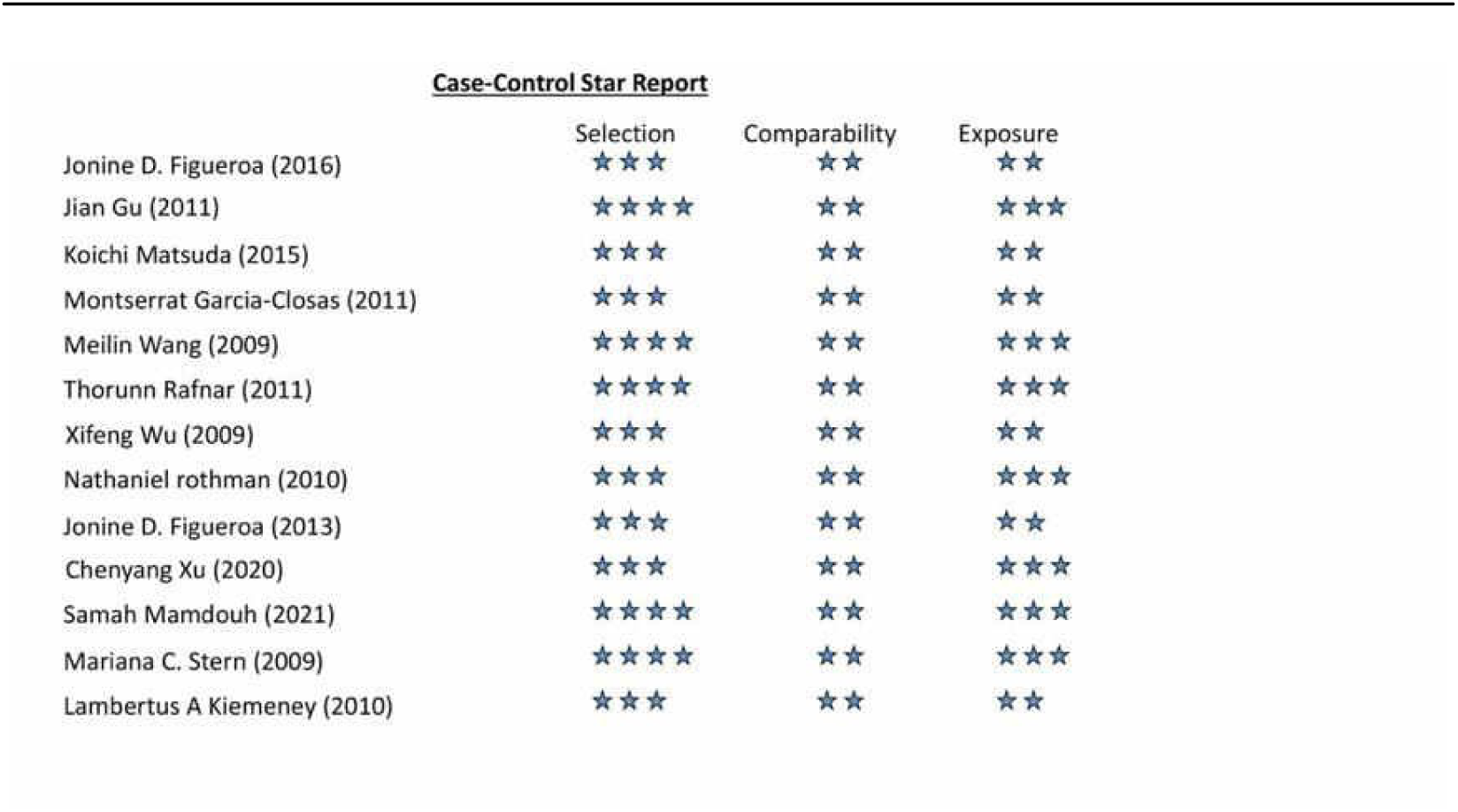
Assessment of Included Prospective Studies’ Bias Risk Using the Newcastle-Ottawa Scale.

### 3.3 Quality assessment

Twelve of the included studies (92%) were rated high quality based on the NOS assessment procedure. These studies had a total of between 8-10 stars (table 4.2) based on subject selection, comparability of the cases and controls, and exposure to experimental procedures. Only one study was rated moderate because of the absence of clear information on the selection and exposure domains (please insert citation for Chenyang Xu 2020). Therefore, all the 13 included studies had moderate to high quality and hence were included in the quantitative synthesis.

## Discussion

This study presents a comprehensive overview of genetic susceptibility to bladder cancer, drawing from a meticulous systematic review of 13 Genome-Wide Association Studies (GWAS). It is worth noting that this systematic review represents one of the most extensive analyses of bladder cancer-related GWAS to date. Our objective was to synthesize the findings of these studies, shedding light on the genetic correlations between bladder cancer and associated susceptible genes. This review was necessitated by the significant public health impact of bladder cancer, which accounted for an estimated 16,000 fatalities in the United States in 2015, affecting both men and women (Wang *et al*., 2016).

### Association between the polymorphisms and bladder cancer risk

Within the spectrum of genetic variations associated with bladder cancer, chromosome 18q12.3 emerges as particularly vulnerable, harboring four polymorphisms on the same locus. These include rs7238033, rs10775480, and rs11082469 on 18q12.3 (Garcia-closas *et al*., 2011), as well as rs17674580 on chromosome 18q12.3 (Xu *et al*., 2020). Additionally, three polymorphisms were reported on chromosome 5p15.3, with rs2736098 and rs401681 on 5p15.33 (Rafnar *et al*., 2011) and rs401681 on 5p15.3 (Xu *et al*., 2020). Among the studies, the largest GWAS, conducted by Rafnar *et al*., (2011), involving 4147 cases and 34988 controls, identified rs9642880 G/A and rs710521 C/T as having stronger associations with bladder cancer risk (P=0.125 and P=0.254, respectively). Figueroa *et al*., (2015) identified rs6104690 and rs6108803 as significantly linked to MICB risk (P ≤ 0.03). Intriguingly, when accounting for the recently discovered SNP rs6108803 in locus 20p12.2, the link for 20p12.2 rs62185668 was no longer significant (P = 0.25) (Figueroa *et al*., 2015).

Furthermore, Matsuda *et al*., (2015) identified that the SNP rs398652 on 14q21, previously associated with leukocyte telomere length, was linked to a lower risk of bladder cancer (P = 0.028771). Chromosome 15q24’s SNP also showed a significant correlation with bladder cancer (P-value of 1.22 × 10^−24) according to Matsuda *et al*., (2015). Wang *et al*., (2009) found that individuals with the GT/TT genotype of the rs9642880 G/T polymorphism had a significantly increased risk of bladder cancer compared to those with the GG genotype. Additionally, a missense mutation (rs2294008) in the PSCA gene consistently correlated with bladder cancer in US and European populations (Wu et al., 2012). Stern *et al*., (2009) suggested that the rs710521 A allele may increase the risk of developing low-risk urinary bladder cancers.

### Association between the genes and bladder cancer risk

The genetic factors implicated in bladder cancer risk are multifaceted. For instance, Pellino homolog 2 (PELI2) was linked to the inflammatory response and cytokine production, suggesting that chronic inflammation could contribute to the relationship between rs398652 telomere length and cancer risk (Gu *et al*., 2011). Higher activity of the CYP1A2 gene on chromosome 15q24, which metabolizes carcinogenic compounds from tobacco, was associated with an increased risk of tobacco-related malignancies (Matsuda *et al*., 2015). The solute carrier family 14 member 1 gene (SLC14A1), governing urine volume and concentration in the kidney, may offer new insights into the etiology of bladder cancer (Garcia-closas *et al*., 2011). MYC, a multifunctional protein controlling cell division, proliferation, and apoptosis, exhibited higher mRNA and protein levels in bladder tissues of individuals with the rs9642880 GT/TT genotypes (Mamdouh *et al*., 2022; Wang *et al*., 2009). In the region of TP63 on chromosome 3q28, Rs710521 and TP53 were identified, with the A allele of rs710521 strongly associated with an increased risk of bladder cancer in the Egyptian population (Wen *et al*., 2019).

## Conclusion

Despite this comprehensive analysis, our understanding of the genetic predisposition to bladder cancer remains incomplete, with the majority of robust data derived from GWAS. This systematic review did not yield additional genetic association evidence.

As the current knowledge of genetic susceptibility to bladder cancer has limited implications for public health, larger cohorts are imperative to identify more significant polymorphisms across the genome. Further research is essential to unravel the complex genetic underpinnings of bladder cancer and pave the way for targeted interventions and personalized treatments.

## Declarations

### Availability of data and material

All data associated with this study are accessible via this link: https://github.com/babasaraki/systematic-review-GWAS

### Authors’ contributions

Conceives and designed the study: UA, Performed literature mining an wrote the first draft: AAA & UM, Review and re-write the draft: UA & BI, Performed the analysis: SHK, Review the draft: UAG & MMJ.

## Data Availability

All data produced are available online at https://github.com/babasaraki/systematic-review-GWAS

https://github.com/babasaraki/systematic-review-GWAS/tree/main

## List of abbreviations

(Bca/BC): Bladder cancer
(GWAS): Genome-wide associated studies
(SNP/SNPs): Single nucleotide polymorphism
(MIBC): Muscular-invasive bladder cancer
(NMIBC): Non-muscular invasive bladder cancer
(UC): Urothelial Carcinoma
(SCC): Squamous cell carcinoma
(TCC): Transitional cell carcinoma

## Reference

1. Bau, D.-T., Chang, C.-H., Tsai, R.-Y., Wang, H.-C., Wang, R.-F., Tsai, C.-W., Yao, C.-H., Chen, Y.-S., Shyue, S.-K., & Huang, C.-Y. (2011). Significant association of caveolin-1 genotypes with bladder cancer susceptibility in Taiwan. The Chinese Journal of Physiology, 54(3), 153–160. 10.4077/cjp.2011.amm009

2. Chang, M., He, L., & Cai, L. (2018). An Overview of Genome-Wide Association Studies. Methods in Molecular Biology (Clifton, N.J.), 1754, 97–108. 10.1007/978-1-4939-7717-8_6

3. Dobruch, J., & Oszczudłowski, M. (2021). Bladder Cancer: Current Challenges and Future Directions. Medicina (Kaunas, Lithuania), 57(8). 10.3390/medicina57080749

4. Figueroa, J. D., Middlebrooks, C. D., Banday, A. R., Ye, Y., Garcia-Closas, M., Chatterjee, N., Koutros, S., Kiemeney, L. A., Rafnar, T., Bishop, T., Furberg, H., Matullo, G., Golka, K., Gago-Dominguez, M., Taylor, J. A., Fletcher, T., Siddiq, A., Cortessis, V. K., Kooperberg, C., … Rothman, N. (2015). Identification of a novel susceptibility locus at 13q34 and refinement of the 20p12.2 region as a multi-signal locus associated with bladder cancer risk in individuals of european ancestry. Human Molecular Genetics, 25(6), 1203–1214. 10.1093/hmg/ddv492

5. Figueroa, J. D., Ye, Y., Siddiq, A., Garcia-closas, M., Chatterjee, N., Prokunina-olsson, L., Cortessis, V. K., Kooperberg, C., Cussenot, O., Benhamou, S., Prescott, J., Porru, S., Dinney, C. P., Malats, N., Baris, D., Purdue, M., Jacobs, E. J., Albanes, D., Wang, Z., … Rothman, N. (2014). Genome-wide association study identifies multiple loci associated with bladder cancer risk. Human Molecular Genetics, 23(5), 1387–1398. 10.1093/hmg/ddt519

6. Garcia-closas, M., Ye, Y., Rothman, N., Figueroa, J. D., Malats, N., Dinney, C. P., Chatterjee, N., Prokunina-olsson, L., Wang, Z., Lin, J., Real, F. X., Jacobs, K. B., Baris, D., Thun, M., De vivo, I., Albanes, D., Purdue, M. P., Kogevinas, M., Kamat, A. M., … Wu, X. (2011). A genome-wide association study of bladder cancer identifies a new susceptibility locus within SLC14A1, a urea transporter gene on chromosome 18q12.3. Human Molecular Genetics, 20(21), 4282–4289. 10.1093/hmg/ddr342

7. Gu, J., Chen, M., Shete, S., Amos, C. I., Kamat, A., Ye, Y., Lin, J., Dinney, C. P., & Wu, X. (2011). A genome-wide association study identifies a locus on chromosome 14q21 as a predictor of leukocyte telomere length and as a marker of susceptibility for bladder cancer. Cancer Prevention Research, 4(4), 514–521. 10.1158/1940-6207.CAPR-11-0063

8. Gupta, S., Rajiah, P., Middlebrooks, E. H., Baruah, D., Carter, B. W., Burton, K. R., Chatterjee, A. R., & Miller, M. M. (2018). Systematic Review of the Literature: Best Practices. Academic Radiology, 25(11), 1481–1490. 10.1016/j.acra.2018.04.025

9. Hamdi, Y., Abdeljaoued-tej, I., Zatchi, A. A., Abdelhak, S., Boubaker, S., Brown, J. S., & Benkahla, A. (2021). Cancer in Africal: The Untold Story. 11(April), 1–19. 10.3389/fonc.2021.650117

10. Kellermeyer, L., Harnke, B., & Knight, S. (2018). Covidence and rayyan. Journal of the Medical Library Association: JMLA, 106(4), 580.

11. Kiemeney, L. A., Sulem, P., Besenbacher, S., Vermeulen, S. H., Gudmundsson, J., Zanon, C., Kostic, J., Masson, G., Bjarnason, H., Grotenhuis, A. J., Verhaegh, G. W., Bishop, D. T., Sak, S. C., Elliott, F., Barrett, J. H., Hurst, C. D., & Verdier, P. J. De. (2010). Europe PMC Funders Group Europe PMC Funders Author Manuscripts A sequence variant at 4p16. 3 confers susceptibility to urinary bladder cancer. 42(5), 415–419. 10.1038/ng.558.A

12. Littell, J., Corcoran, J., & Pillai, V. (2009). Systematic Reviews and Meta-Analysis. In Journal of Pain - J PAIN (Vol. 5). 10.1093/acprof:oso/9780195326543.001.0001

13. Lo, C. K.-L., Mertz, D., & Loeb, M. (2014). Newcastle-Ottawa Scale: comparing reviewers’ to authors’ assessments. BMC Medical Research Methodology, 14, 45. 10.1186/1471-2288-14-45

14. Mamdouh, S., Khorshed, F., Hammad, G., Elesaily, K., Safwat, G., Hammam, O., & Aboushousha, T. (2022). Molecular Detection of Genetic Susceptibility to Bladder Cancer in Egyptian Patients. Asian Pacific Journal of Cancer Prevention, 23(1), 221–232. 10.31557/APJCP.2022.23.1.221

15. Matsuda, K., Takahashi, A., Middlebrooks, C. D., Obara, W., Nasu, Y., Inoue, K., Tamura, K., Yamasaki, I., Naya, Y., Tanikawa, C., Cui, R., Figueroa, J. D., Silverman, D. T., Rothman, N., Namiki, M., Tomita, Y., Nishiyama, H., Kohri, K., Deguchi, T., … Shuin, T. (2015). Genome-wide association study identified SNP on 15q24 associated with bladder cancer risk in Japanese population. Human Molecular Genetics, 24(4), 1177–1184. 10.1093/hmg/ddu512

16. Nahar, K. N., Nessa, A., Shamim, S., Nasrin, B., Hossain, F., & Begum, N. (2011). Role of VIA in cervical cancer screening in low-resource countries. Mymensingh Medical Journall: MMJ, 20(3), 528–535.

17. Rafnar, T., Vermeulen, S. H., Sulem, P., Thorleifsson, G., Aben, K. K., Alfredwitjes, J., Grotenhuis, A. J., Verhaegh, G. W., Kaa, C. A. H. De Besenbacher, S., Gudbjartsson, D., Stacey, S. N., Gudmundsson, J., Johannsdottir, H., Bjarnason, H., Zanon, C., Helgadottir, H., Jonasson, J. G., Tryggvadottir, L., … Stefansson, K. (2011). European genome-wide association study identifies SLC14A1 as a new urinary bladder cancer susceptibility gene. 20(21), 4268–4281. 10.1093/hmg/ddr303

18. Richters, A., Aben, K. K. H., & Kiemeney, L. A. L. M. (2020). The global burden of urinary bladder cancer: an update. World Journal of Urology, 38(8), 1895–1904. 10.1007/s00345-019-02984-4

19. Selvaraj, N., Dholakia, K., & Ragavan, N. (2021). A Single Tertiary Center Experience in a South Asian Population: Does Tobacco Use Influence Bladder Cancer? Cureus, 13(10), e18734. 10.7759/cureus.18734

20. Shamseer, L., Moher, D., Clarke, M., Ghersi, D., Liberati, A., Petticrew, M., Shekelle, P., & Stewart, L. A. (2015). Preferred reporting items for systematic review and meta-analysis protocols (PRISMA-P) 2015: elaboration and explanation. BMJ (Clinical Research Ed.), 350, g7647. 10.1136/bmj.g7647

21. Siddaway, A. P., Wood, A. M., & Hedges, L. V. (2019). How to Do a Systematic Review: A Best Practice Guide for Conducting and Reporting Narrative Reviews, Meta-Analyses, and Meta-Syntheses. Annual Review of Psychology, 70, 747–770. 10.1146/annurev-psych-010418-102803

22. Siracusano, S., Rizzetto, R., & Porcaro, A. B. (2020). Bladder cancer genomics. Urologia, 87(2), 49–56. 10.1177/0391560319899011

23. Stern, M. C., Van Den Berg, D., Yuan, J. M., Conti, D. V., Gago-Dominguez, M., Pike, M. C., Xiang, Y. B., Gao, Y. T., & Cortessis, V. K. (2009). Sequence variant on 3q28 and urinary bladder cancer risk: Findings from the Los Angeles-Shanghai bladder case-control study. Cancer Epidemiology Biomarkers and Prevention, 18(11), 3057–3061. 10.1158/1055-9965.EPI-09-0492

24. Verma, M. (2016). Genome-wide association studies and epigenome-wide association studies go together in cancer control. 12, 1645–1664.

25. Wang, M., Li, Z., Chu, H., Lv, Q., Ye, D., Ding, Q., Xu, C., Guo, J., Du, M., Chen, J., Song, Z., Yin, C., Qin, C., Gu, C., Zhu, Y., & Xia, G. (2016). Genome-Wide Association Study of Bladder Cancer in a Chinese Cohort Reveals a New Susceptibility Locus at 5q12. 3. 3277–3285. 10.1158/0008-5472.CAN-15-2564

26. Wang, M., Wang, M., Zhang, W., Yuan, L., Fu, G., Wei, Q., & Zhang, Z. (2009). Common genetic variants on 8q24 contribute to susceptibility to bladder cancer in a Chinese population. Carcinogenesis, 30(6), 991–996. 10.1093/carcin/bgp091

27. Wu, X., Ye, Y., Kiemeney, L. a, Sulem, P., Rafnar, T., Matullo, G., Seminara, D., Yoshida, T., Saeki, N., & Angeline, S. (2012). Confers Susceptibility To Urinary Bladder Cancer. 41(9), 991–995. 10.1038/ng.421.Genetic

28. Xu, C., Lin, X., Qian, W., Na, R., Yu, H., Jia, H., Jiang, H., Fang, Z., Zheng, S. L., Ding, Q., Wu, Y., Zheng, J., & Xu, J. (2020). Genetic risk scores based on risk-associated single nucleotide polymorphisms can reveal inherited risk of bladder cancer in Chinese population. Medicine (United States), 99(19). 10.1097/MD.0000000000019980

29. Ye, Y., Rothman, N., Figueroa, J. D., Wang, Z., Lin, J., Real, F. X., Jacobs, K. B., Baris, D., Thun, M., Vivo, I. De, Albanes, D., Purdue, M. P., Kogevinas, M., Kamat, A. M., Lerner, S. P., Grossman, H. B., Johnson, A., Schwenn, M., Karagas, M. R., … Wu, X. (2011). A genome-wide association study of bladder cancer identifies a new susceptibility locus within SLC14A1, a urea transporter gene. 20(21), 4282–4289. 10.1093/hmg/ddr342

