## Supplementary Table 1 for "A systematic review of genome-wide association studies on bladder cancer"

**Supplementary 1:** Search terms and strategies

1. **PubMed**: a medical subject subheadings (MeSH) term were generated for search in the PubMed database; ("Genome-Wide associated Study/classification"[Mesh] OR "Genome-Wide associated Study/ethics"[Mesh] OR "Genome-Wide associated Study/history"[Mesh] OR "Genome-Wide associated Study/instrumentation"[Mesh] OR "Genome-Wide associated Study/methods"[Mesh] OR "Genome-Wide associated Study/standards"[Mesh] OR "Genome-Wide associated Study/statistics and numerical data"[Mesh] OR "Genome-Wide associated Study/trends"[Mesh]) OR ( "Urinary Bladder Neoplasms/analysis"[Mesh] OR "Urinary Bladder Neoplasms/anatomy and histology"[Mesh] OR "Urinary Bladder Neoplasms/classification"[Mesh] OR "Urinary Bladder Neoplasms/complications"[Mesh] OR "Urinary Bladder Neoplasms/congenital"[Mesh] OR "Urinary Bladder Neoplasms/cytology"[Mesh] OR "Urinary Bladder Neoplasms/diagnosis"[Mesh] OR "Urinary Bladder Neoplasms/diagnostic imaging"[Mesh] OR "Urinary Bladder Neoplasms/diet therapy"[Mesh] OR "Urinary Bladder Neoplasms/drug therapy"[Mesh] OR "Urinary Bladder Neoplasms/embryology"[Mesh] OR "Urinary Bladder Neoplasms/epidemiology"[Mesh] OR "Urinary Bladder Neoplasms/ethnology"[Mesh] OR "Urinary Bladder Neoplasms/aetiology"[Mesh] OR "Urinary Bladder Neoplasms/genetics"[Mesh] OR "Urinary Bladder Neoplasms/history"[Mesh] OR "Urinary Bladder Neoplasms/immunology"[Mesh] OR "Urinary Bladder Neoplasms/mortality"[Mesh] OR "Urinary Bladder Neoplasms/organization and administration"[Mesh] OR "Urinary Bladder Neoplasms/pathology"[Mesh] OR "Urinary Bladder Neoplasms/physiology"[Mesh] OR "Urinary Bladder Neoplasms/physiopathology"[Mesh] OR "Urinary Bladder Neoplasms/prevention and control"[Mesh] OR "Urinary Bladder Neoplasms/psychology"[Mesh] OR "Urinary Bladder Neoplasms/radiotherapy"[Mesh] OR "Urinary Bladder Neoplasms/rehabilitation"[Mesh] OR "Urinary Bladder Neoplasms/secondary"[Mesh] OR "Urinary Bladder Neoplasms/statistics and numerical data"[Mesh] OR "Urinary Bladder Neoplasms/surgery"[Mesh] OR "Urinary Bladder Neoplasms/therapy"[Mesh] OR "Urinary Bladder Neoplasms/ultrastructure"[Mesh] OR "Urinary Bladder Neoplasms/urine"[Mesh] OR "Urinary Bladder Neoplasms/veterinary"[Mesh] OR "Urinary Bladder Neoplasms/virology"[Mesh] ).
2. **Google scholar**: ((((((genome wide associated studies) OR (GWAS)) OR (Genome wide associated analysis)) AND (Bladder)) AND (cancer)) OR (carcinoma)) OR (neoplasms)
3. **Cochrane library**: (GWAS):ti,ab,kw OR (genome wide associated studies):ti,ab,kw AND (Bladder cancer):ti,ab,kw OR ("bladder carcinoma"):ti,ab,kw AND (Bladder neoplasms):ti,ab,kw (Word variations have been searched)	829.
